# Chronic Pain and Eye Movements: A NeuroIS Approach to Designing Smart Clinical Decision Support Systems

**DOI:** 10.1101/2023.08.04.23293108

**Authors:** Doaa Alrefaei, Soussan Djamasbi, Diane Strong

## Abstract

The pressing need for objective measures in the evaluation of chronic pain both in research and practice highlights the role that neuro information systems (NeuorIS) research plays in designing smart clinical decision support systems. A first step in such a research agenda is identifying practical stimuli-task paradigms that can reliably detect chronic pain from physiological measures such as eye movements. In this study, we propose and test a new stimuli-task paradigm. Our results show that our proposed stimuli-task paradigm can detect differences in information processing behavior of people with and without chronic pain. The results also show that our proposed stimuli-task paradigm can reliably predict a person’s reported subjective pain experience from his/her eye movements. These findings provide support for our proposed stimuli-task paradigm. They also show that the eye-tracking variables that we selected to test our proposed paradigm are effective in capturing the impact of chronic pain on visual attention and suggest that eye movements have the potential to serve as reliable biomarkers of chronic pain. In other words, our results provide support for the potential of eye movements to facilitate the development of smart information systems that can detect the presence and/or the severity of chronic pain from an individual’s ocular behavior.

## Introduction

As more and more human needs are addressed by digital goods and services, competition in today’s digital economy is driven by continual market demand for innovation (Djamasbi and Strong, 2019). One way to address this market demand is by developing smart devices that can provide useful services with outstanding user experiences. Advances in technology provide us with the opportunity to collect physiological measures, such as eye movements, unobtrusively without any burden on users. Hence, neuro Information system (NeuroIS) research, which uses physiological measures to detect changes in user experience and/or behavior (Bačić and Henry, 2022; Mirhoseini et al., 2022; Loos et al., 2022; Jia et al., 2021), plays an increasingly critical role in designing smart products (Fehrenbacher and Djamasbi, 2017; Shojaeizadeh et al., 2019).

One domain that can benefit from developing smart products that use sensor-based technologies is chronic pain, which refers to “a distressing experience associated with actual or potential tissue damage” that lasts more than three months (Crofford, 2015; Merskey, 1986). As one of the most common chronic conditions (CDC, 2020; Young et al., 2022), the persistent experience of pain is a major health problem with severe personal, societal, and economic negative consequences (CDC, 2020; Phillips, 2006; Yong et al., 2022).

Providing effective treatment for chronic pain requires a comprehensive understanding of the pain experience. Chronic pain is typically assessed by collecting self-reported ratings that capture a patient’s level of pain intensity as well as the degree to which pain interferes with the patient’s physical and day-to-day activities (McCahon et al., 2005; Rose et al., 2018). While self-reported ratings provide an opportunity for individuals to convey their point of view, they lack the objectivity that is needed to have a more balanced (objective and subjective) view of pain experience (Borsook et al., 2011). Because objectivity in chronic pain assessment can provide major improvements in the effective treatment of chronic pain (Borsook et al., 2011; Xu and Huang, 2020), a NeuroIS research agenda focusing on developing smart information systems that can detect chronic pain objectively, via physiological measures, is of great importance to both research and practice. Our study provides a first step in such a research agenda.

NeuroIS literature shows that eye movements can reliably measure and detect changes in a person’s information processing and decision behavior (Fehrenbacher and Djamasbi, 2017; Shojaeizadeh et al., 2019). Because chronic pain impacts attention (Phelps et al., 2021), and eye movements provide moment-to-moment information about changes in attention, it is reasonable to argue that eye movements may serve as excellent physiological measures for detecting chronic pain. For example, smart clinician support systems that could create chronic pain reports from objective eye movements would be useful to clinicians in gaining a more comprehensive understanding of their patients’ chronic pain experience. This argument is supported by a number of eye-tracking studies that show eye movements can capture objective information-rich differences in visual attention to pain stimuli between people with and without chronic pain (e.g., Alrefaei et al., 2022; Fashler and Katz, 2016; Gaffiero et al., 2019).

Developing an effective stimuli-task paradigm is a fundamental step in the NeuroIS research agenda that focuses on detecting chronic pain from eye movement behavior. A recent systematic review of chronic pain studies suggests that the commonly used stimuli-task paradigm may not be suitable for designing smart NeuroIS for detecting chronic pain from eye movement data (Chan et al., 2020). According to this review article, the stimuli-task paradigm that is widely used to examine the impact of chronic pain on visual attention produces mixed results (Chan et al., 2020). One reason could be that the commonly used stimuli-task paradigm does not offer enough opportunities for capturing the complex and dynamic nature of attention because it engages people in relatively simple cognitive activities that are completed in fixed short time intervals.

Hence, in our study, we propose and test a new stimuli-task paradigm that naturally provides more chances for capturing nuances in visual attention. Grounded in theories that explain the interruptive function of pain on information processing (Eccleston and Crombez, 1999; Todd et al. 2015), we argue that engaging people in completing pain-related surveys provides a sensitive stimuli-task paradigm for detecting differences in visual information processing behavior of people with and without chronic pain. Because completing surveys engages people in a more complex cognitive activity than the tasks currently used in chronic pain studies, our proposed stimuli-task paradigm offers more opportunities to observe the impact of chronic pain on cognition. Furthermore, our proposed stimuli-task paradigm naturally increases the prospect of observing the interruptive function of pain on attention because it does not impose a time limit. By using the same surveys that clinicians use to collect subjective pain measures to evaluate chronic pain, our proposed stimuli-task paradigm represents a practical and ecologically valid choice for the NeuroIS research agenda that aims at developing feasible solutions for detecting chronic pain from visual information processing behavior.

## Background

### Eye Tracking to Detect Visual Behavior

Video-based eye-tracking has become increasingly popular in NeuroIS research due to its ability to unobtrusively measure visual behavior (Djamasbi, 2014). Video-based eye-trackers capture a person’s gaze on a visual display by recording and measuring the changes in the person’s pupil position at any given time. The eye-tracking device, which is typically mounted beneath the visual display (e.g., the computer monitor that presents the stimuli), shines invisible infrared light onto the person’s eyes. The reflection of this light, which produces a small bright light on the eye surface (glint) and makes the detection of the pupil easier, is captured by the infrared (IR) sensing video camera embedded in the eye-tracking device. Using the relative position of the glint and pupil center, the eye tracking software can calculate a person’s gaze point on the stimulus (Nyström et al., 2013). Video-based eye trackers capture gaze data continuously with high sampling rates (e.g., 60 to 600 HZ), hence, they provide an excellent tool for capturing the dynamic nature of attention.

The raw gaze stream collected by eye trackers is typically processed to identify gaze points that form fixations and saccades. Fixations refer to a group of stable gaze points that are near in both spatial and temporal proximity. Grounded in the “eye-mind” assumption (Just and Carpenter 1980) it is widely agreed that such clusters of gaze points serve as a reliable indicator of attention to stimuli (Djamasbi, 2014; Rosch and Vogel-Walcutt, 2013; Poole and Ball, 2005). Saccades refer to small, rapid gaze points that occur between fixations (Goldberg and Kotval, 1999). While visual information is not processed during saccadic eye movements, they provide valuable information about how people shift their attention from one focal point to another (Holmqvist et al., 2001; Jacob & Karn, 2003).

### Chronic Pain and Attentional Bias

Chronic pain is a complex and debilitating phenomenon that exerts a significant impact on memory. Those who endure chronic pain often encounter difficulties in suppressing or eliminating painful memories. This cognitive process may culminate in heightened selective attention toward pain-related information (Phelps et al., 2021; Todd et al., 2015). Such selective attention is called ***Attentional Bias*** and refers to the tendency to pay attention selectively to information that is related to one’s concern (Keogh et al., 2001).

Attentional bias toward stimuli that are pain-related can manifest as an increased vigilance toward painful stimuli, or a decreased ability to disengage from such stimuli (Keogh et al., 2001). However, individuals with chronic pain do not always demonstrate their tendency towards selective attention (i.e., attentional bias) by demonstrating heightened focus on pain-related stimuli. In some instances, studies indicate individuals with chronic pain exhibit attentional bias by avoiding pain-related stimuli rather than by paying extra attention to them (Chan et al., 2020; Fashler and Katz, 2016, 2014; Gaffiero et al., 2019; Vervoort et al., 2013).

The observation of diametrically opposing attentional biases (heightened attention to vs. avoidance of pain stimuli) in chronic pain studies can be elucidated through theories that seek to explain the interruptive function of pain with regard to attentional information processing (Eccleston and Crombez, 1999; Todd et al., 2015). These theories suggest that the impact of pain on cognition is influenced by pain-related characteristics such as pain intensity. When an individual experiences severe chronic pain, the appraisal process of their pain experience is more likely to result in one’s heightened attention toward the pain. In contrast, when the chronic pain experience is less severe, the appraisal process may prompt individuals to seek any type of interruption, such as shifting attention from the pain to a task at hand, as a distraction strategy to escape the pain (Eccleston and Crombez, 1999).

### Stimuli-Task Paradigm

Stimuli-task paradigm refers to the context within which behavioral data is collected. Stimuli refers to experimental materials that are prepared for a study (e.g., webpages, dashboards). Task refers to the activity involving the stimuli that participants are required to do in a study (e.g., retrieving information, making decisions). Naturally, the stimuli-task paradigm plays a significant role in behavioral research, particularly in the NeuroIS research that relies on eye-tracking sensors. This is because in addition to providing the context for understanding user behavior (e.g., perceptions and evaluations), the mere arrangement of content on visual stimuli (e.g., structure and format) as well as what people are asked to do with the visual stimuli (e.g., review the content or look for specific information) has a major impact on eye-movement patterns with which the presented information is processed (e.g., Djamasbi and Hall-Philips, 2014; Djamasbi et al., 2009; Cyr et al., 2009).

#### Stimuli-Task Paradigm in Chronic Pain Literature

The stimuli-task paradigm in chronic pain literature is designed to examine differences in attentional bias toward pain-related information. The most commonly used stimuli-task paradigm in this literature is the stimuli presentation paradigm using the dot-probe task (Cardoso et al. 2021), which predominantly relies on the measurement of reaction time to visual probes (Asmundson et al., 2005; Fashler & Katz, 2014, 2016; Franklin et al., 2019; Keogh et al., 2001; Vervoort et al., 2013). The reaction time to visual probes is typically determined by presenting participants with a pair of cues displayed laterally on a screen (e.g., words or images that are placed on the right and left sides of the screen). After a short period of time (e.g., 1500-2500 milliseconds) a new screen is presented to viewers. This screen displays a single dot in the location of one of the two cues in the prior screen. Participants are then required to identify the location of the dot on the screen (e.g., left, or right side of the screen) as soon as possible typically by using a keyboard press or a mouse click. The reaction time (the amount of time that it takes a participant to recognize the location of the dot) is then used to measure attention to the cue on previous screen that was presented in the same location as the dot (Figure 1). The faster the reaction time the more heightened the attention to the cue that was replaced by the dot.

**Figure 1.**
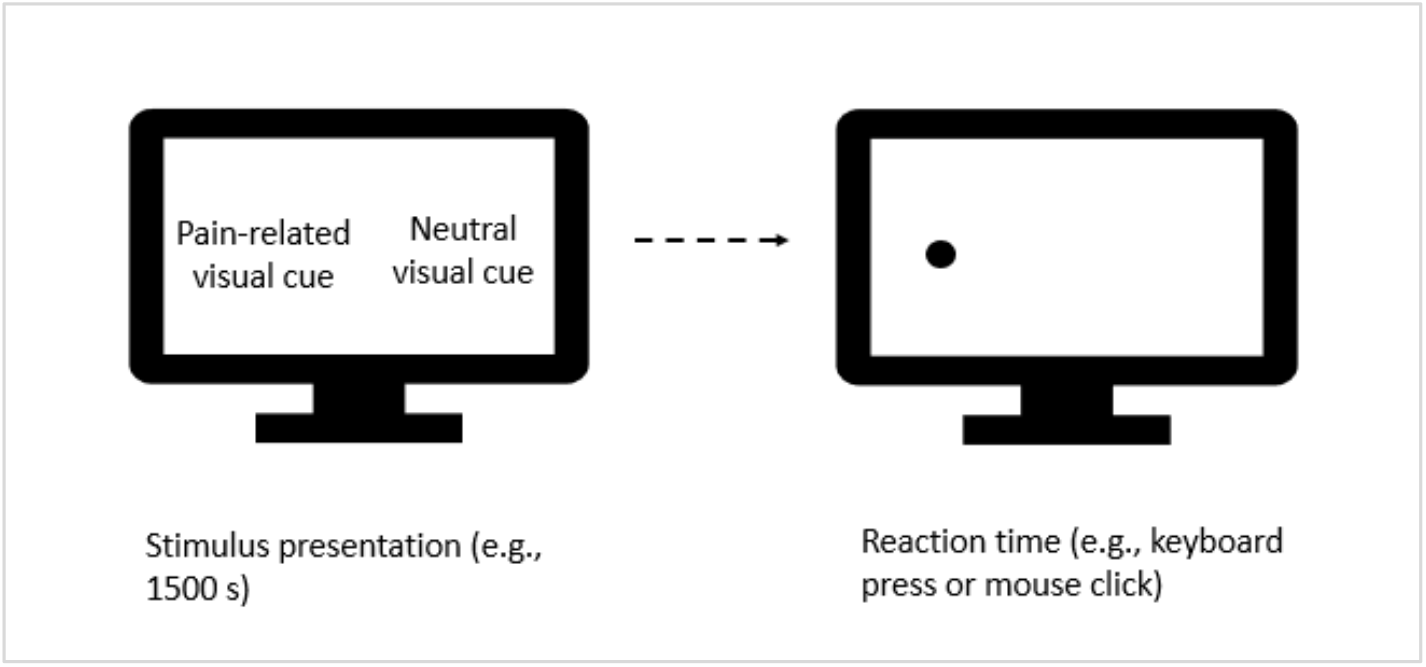
Measuring attention with reaction time in a dot-probe task. Faster reaction times indicate heightened attention to the visual cue that was replaced by the dot.

More recently, eye tracking is used in free viewing tasks (i.e., presenting participants with a set of images in short time intervals) as well as dot-probe tasks to directly measure attention by tracking visual engagement with or shift of focus away from stimuli (Fashler and Katz, 2014, 2016; Gaffiero et al., 2019; Mahmoodi-Aghdam et al. 2017; Vervoort et al., 2013; Yang et al., 2012). In addition to serving as a direct measure of visual attention, moment-to-moment eye movements are likely to provide a more accurate representation of the dynamic nature of attention in chronic pain research (Crombez et al., 2015). The value of eye tracking in chronic pain literature is demonstrated by studies that capture attention via both reaction time and eye tracking. While these studies find no significant differences in reaction time between chronic pain individuals and controls, they show significant differences in eye movement behavior between the two groups (Thigpen et al., 2018).

Despite being touted as the gold standard for capturing attentional bias (Gaffiero et al., 2019), a recent systematic literature review (SLR) of studies that use eye tracking to examine pain-related attentional processes (Chan et al., 2020) shows that using gaze behavior to investigate the impact of chronic pain on information processing behavior is still in its infancy. For example, the SLR article (Chan et al. 2020) resulted in only 24 papers. Of these 24 papers, only 11 studies investigated the impact of chronic pain on attentional processes. Of the 11 articles that examined the impact of chronic pain, only 9 studies examined the interruptive function of chronic pain on attention by comparing the ocular behavior of people with and without chronic pain. Because our study focuses on comparing the viewing behavior of those who suffer from chronic pain and those who are pain free, we summarized the findings of the 9 aforementioned reviewed articles in Table 7 (see Appendix A). The complete listing of all the reviewed articles as well as their details (e.g., methodology, results) can be found in Table A1 of the SLR article (Chan et al. 2020, pp. 5-10).

Table 7 shows that the comparison of gaze behavior between people with and without chronic pain has produced mixed findings. A study’s ability to effectively detect differences in viewing behavior of people with and without chronic pain depends largely on how successfully the stimuli-task paradigm can capture the nuances of visual behavior that are caused by the interruptive function of pain on attention (Chan et al., 2020). All the articles displayed in Table 7 utilize the commonly used stimuli presentation paradigm in pain literature. Hence, the reported results in Table 7 indicate that the commonly used stimuli presentation paradigm in pain literature has not been always successful in detecting differences in viewing behavior between people with and without chronic pain (see Appendix A).

One reason could be that this paradigm cues participants indirectly to think about their pain experience. This approach may make the stimuli-task paradigm less personally relevant to participants (Chan et al., 2020). Another reason could be the short, fixed exposure times used in this paradigm. All reviewed studies summarized in Table 7 used fixed short periods of time (e.g., 500-4000 milliseconds) to capture attentional bias toward visual stimuli. Given the complex and dynamic nature of attention, such fixed short times provide only a narrow window for capturing important nuances in information processing behavior. The mixed results could also be due to relatively simple visual stimuli (e.g., single words and/or images) and relatively simple tasks (e.g., viewing the stimuli and/or reporting whether a dot appears on the left or right side of the screen) used in the stimuli presentation paradigm.

#### Our Proposed Stimuli-Task Paradigm

We conjecture that a more context-rich visual stimulus and a more demanding task are likely to provide more opportunities for capturing the interruptive function of chronic pain on information processing behavior. Our conjecture is supported by a recent eye-tracking study that uses pain-related surveys as visual stimuli and the process of completing the surveys (i.e., assessing one’s pain-related health symptoms) as the experimental task. This study shows significant differences between people with and without chronic pain in the number of times they attended to survey option labels (e.g., *Not at all*, *A little bit*, *Somewhat, Quite a bit*, *Very much*) (Alrefaei et al., 2022). The results of this recent eye-tracking study suggest that completing pain-related surveys may serve as a powerful stimuli-task paradigm for capturing nuances in ocular behavior that represent attentional bias. Such a stimuli-task paradigm is likely to be perceived as personally relevant because it engages people directly in appraising their health symptoms. It provides a context-rich environment for making decisions (i.e., choosing an option among a set of alternatives that best reflects one’s experience of a health symptom) without imposing fixed short time limits. The rich decision-making context of such stimuli-task paradigm along with constraint-free exposure time, naturally provides more opportunities for capturing the dynamic nature of attention.

Hence, we hypothesize that:

*H1. Completing pain-related surveys provides a sensitive stimuli-task paradigm for capturing differences in attentional bias between people with and without chronic pain*.

The above-stated hypothesis examines the sensitivity of pain-related survey stimuli-task paradigm in detecting presence/absence of chronic pain.

Our next hypothesis extends the above examination by testing whether this stimuli-task paradigm is sensitive enough to predict subjective pain experience from objective eye movement data. Our reasoning is grounded in the pain literature that asserts the interruptive nature of chronic pain on information processing behavior depends on the severity of pain experience (Eccleston and Crombez, 1999; Moriarty, McGuire, and Finn, 2011; Todd et al., 2015). Hence, we argue that our proposed stimuli-task paradigm is both effective in detecting the presence/absence of chronic pain (see H1) and effective in revealing the degree to which people experience pain from their objective eye movements. We assert that:

*H2. Completing pain-related surveys provides a sensitive stimuli-task paradigm for predicting subjective pain intensity scores from objective eye movements*.

## Methodology

To test our hypotheses, we conducted an IRB-approved eye-tracking experiment. In the following sections, we explain the methodology that we used to test our hypotheses. We provide details for the visual stimuli and task used in our study. We also explain the process by which we recruited participants for our study and collected experimental data. Then, we provide the specification for the eye-tracker used in our study and discuss the variables that we used to capture visual information processing behavior. We explain how we organized the collected data into chronic-pain and pain-free datasets and discuss how we analyzed the data to test our hypothesis.

### Visual Stimuli and Task

We used 3 pain-related measures (***pain interference***, ***physical function***, and ***pain intensity***) from The Patient-Reported Outcomes Measurement Information System (PROMIS) 29+ v2 profile. PROMIS measures are devised through a concerted effort by the National Institutes of Health (NIH) to provide healthcare professionals and researchers with a standardized national resource for monitoring and evaluating well-being (Cella et al., 2010; Rose et al., 2018). PROMIS measures are designed to evaluate a battery of symptoms across diverse health domains, including physical function, anxiety, fatigue, depression, cognitive function, ability to participate in social roles, sleep disturbance, pain interference, and pain intensity. Because PROMIS measures are commonly used in clinical settings as the conventional means of assessing chronic pain, they provide a practical stimuli-task paradigm for detecting differences in information processing behavior of those who suffer from chronic pain and those who are pain free.

The task in our study required participants to complete the three pain-related measures in PROMIS 29+ v2 profile that we used as visual stimuli, i.e., *pain interference, physical function, and pain intensity.* The *pain interference* measure is used to capture the degree to which pain has interrupted one’s daily activities in the past seven days using four questions: 1) “*How much did pain interfere with your day to day activities?”*, 2) *“How much did pain interfere with work around the home?”*, 3)*“How much did pain interfere with your ability to participate in social activities?”*, and 4) *“How much did pain interfere with your household chores?”*. The *Physical Function* measure is designed to assess the degree to which pain restricts one’s physical activities via four questions: 1) *“Are you able to do chores such as vacuuming or yard work?”*, 2) *“Are you able to go up and down stairs at a normal pace?”*, 3) *“Are you able to go for a walk of at least 15 minutes?”*, and 4) *Are you able to run errands and shop?”*. The *pain intensity* measure is designed to assess the severity of pain experience in the past seven days by asking a single question: “*How would you rate your pain on average?”*. The questions in *pain interference* and *physical function* measures are rated on a 5-point scale. Higher rating values for *pain interference* and *physical function* measures indicate more intense symptoms. The *pain intensity* measure uses an 11-point scale ranging from 0–10. On this scale, a rating of 0 indicates the absence of pain; a rating of 10 indicates the worst pain imaginable.

### Participant Recruitment and Data Collection Process

Using a flyer placed on various community spaces within and outside a northeastern U.S. university, forty-eight adults were recruited to participate in our study (Table 1). Those who were interested in participating in our study were required to answer a screening question about their chronic pain health status. Based on the provided definition for chronic pain, i.e., an intense pain experience (4 or higher on a 0–10 Likert scale) that persists at least for 3 months, the screening question required participants to self-identify as 1) someone with chronic pain, 2) someone who is pain free, 3) or someone with a pain experience that is somewhat between the two previous conditions.

**Table 1:** Participant Demographics.

| Table 1: Participant Demographics |  |
| --- | --- |
| Number of participants | 47; Forty-eight participants were recruited; the data for one was eliminated due to calibration issues |
| Gender | male (n=20), female (n=26), other (n=1) |
| Age range | 18 to 65 (mean=25.60 years, SD= 12.49) |
| Chronic pain status | Chronic pain (n=21), pain free (n=22), in-between (n=4) |
| Chronic pain types | Back pain (n=7/21), neck pain (n=4/21), knee pain (n=4/21), leg pain (n=3/21), headache (n=2/21), shoulder pain (n=2/21), eye pain (n=1/21), muscle pain (n=1/21), and wrist pain (n=1/21). Some participants reported to experience multiple chronic pain conditions; some reported to have been medically diagnosed as chronic pain patients |

Participants were then scheduled for individual experimental sessions at the university laboratory. The data collection process took place over twelve weeks. Before presenting the stimuli (pain-related surveys), the eye tracker was calibrated for each participant. This calibration process took less than a minute. Participants’ eye movements were then collected when they were completing the experimental task (completing the surveys). After completing the task, participants took part in an exit interview. During this interview, the participants’ health status was verified. Once again participants were provided with the definition of chronic pain and were asked to self-identify their pain status based on three categories: suffering from chronic pain, being pain free, or having a health status between chronic pain and being pain free. At the end of the experimental session, each participant was provided with a $20 Amazon gift card.

We were not able to calibrate the eye tracker for one participant. This is consistent with prior studies reporting that a small number of participants may not be able to complete the calibration process successfully (Fehrenbacher and Djamasbi, 2017).

Among the chronic pain participants in our study, a diverse range of pain types was reported. The prevalent types of pain reported were back pain, neck pain, and knee pain. Additionally, other forms of pain, such as leg pain, headache, shoulder pain, eye pain, muscle pain, and wrist pain were also reported. It is important to note that some participants experienced multiple chronic pain conditions, and some were medically diagnosed with at least one chronic pain condition (Table 1).

### Apparatus

Tobii Pro Spectrum 600 Hz was used to collect participants’ eye movements in our study. Participants’ raw gaze data was processed with the IVT filter provided by Tobii Pro Lab software version 1.162.32461 (x64). The threshold for the IVT filter was set to 30°/s. The minimum duration for fixations was set to 100 ms (Liu et al., 2021). Visual stimuli were presented to participants via a desktop computer. Participants’ eye movements were captured unobtrusively during task completion by an eye-tracking machine attached to the monitor with a resolution of 1920X1080 pixels and a screen size of 23.8 inches.

### Eye-Tracking Metrics

We used three eye-movement metrics that have been used in pain literature to capture attention: fixation duration, saccade frequency, and visit duration (Fashler and Katz, 2014, 2016; Skaramagkas et al., 2023; Yang et al., 2012). ***Fixation duration (FD)*** refers to the amount of time when the eye remains relatively still to process visual information. ***Saccade frequency (SF)*** captures the number of times there is a change in focus of attention when a person looks at a stimulus. ***Visit duration (VD)*** refers to the total amount of time that people view the stimulus including the time they spend engaging with the content (via fixations) and the time they spend changing their focus (via saccades).

These three eye-movement metrics were captured for multiple areas, as customary in eye-tracking research (Djamasbi, 2014). We divided each visual stimulus (i.e., *pain interference*, *physical function*, and *pain intensity*) into two major areas of investigation (AOIs) that were relevant to our study. One of these AOIs delineated the part of the stimulus that contained the questions in the pain-related surveys (***Question AOI***). The other AOI covered the part of the stimulus that contained possible answers to those questions (***Answer AOI***). The amount of time that users take to visit an AOI is a suitable measure of attention to that AOI (Djamasbi, 2014). An AOI visit includes both fixation gaze points which represent ***engagement*** with the content of the AOI and saccadic gaze points which represent ***shift in focus*** within that AOI. Both engagement and shift in focus have been widely used in the eye-tracking pain literature to examine attentional bias toward stimuli (Fashler and Katz 2014, 2016; Skaramagkas et al., 2023; Yang et al., 2012).

Because the stimuli-task paradigm in our study did not use a fixed time limit, we normalized the eye-tracking variables used in our study (Shojaeizadeh et al., 2019). To do so, we defined a third AOI that contained the entire screen (Screen AOI). We then calculated fixation duration, saccade frequency, and visit duration for Question and Answer AOIs as the percentage of their total values captured in Screen AOI. For example, the ratio of fixation duration in question AOI was determined by calculating FD-Question-AOI / FD-Screen-AOI, the ratio of saccade frequency was determined by calculating SF-Question-AOI / SF-Screen-AOI, and the ratio of visit duration was determined by calculating VD-Question-AOI / VD-Screen-AOI (Figure 2).

**Figure 2.**
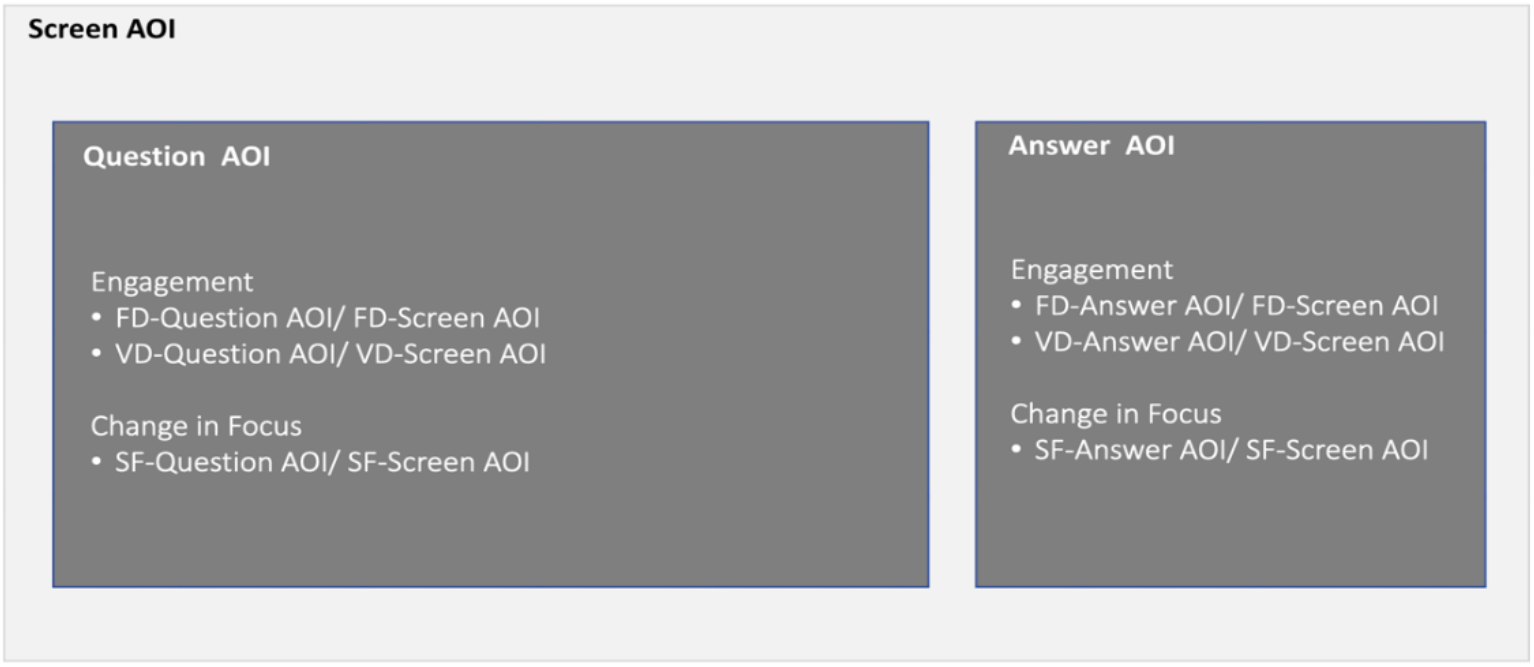
Areas of investigation (AOI) and eye tracking data calculated for each AOI, FD= fixation duration, VD= visit duration, SF= saccade frequency

### Datasets

The eye-tracking dataset included the participant’s eye-movement data for the Question and Answer AOIs on each of the three PROMIS measures that were used as visual stimuli (i.e., 6 AOIs per participant). In addition to eye-movement behavior, the dataset for each participant also included the participant’s self-reported scores for those three PROMIS pain measures. The collected datasets were then organized in chronic-pain (n=21), pain-free (n=22), and in-between (n=4) groups based on participants’ self-identified health status which was solicited at the time they registered for participation and was confirmed during the exit interview portion of the experiment. To examine the impact of chronic pain on eye movements that represent attentional bias, we analyzed only the datasets for those who self-identified as having chronic pain and those who declared to be pain-free (n=43). As shown in Table 2, this process resulted in a total of 258 eye-movement datasets (43 participants * 6 AOI).

**Table 2:** Eye movement dataset.

| Table 2: Eye movement dataset |  |
| --- | --- |
| Number of visual stimuli (3): | Pain interference, physical function, and pain intensity |
| Number of AOIs on each stimulus (2): | Question AOI and Answer AOI |
| Number of eye-movement datasets (258): | 43 participants (21 chronic pain, 22 pain free) * 6 AOIs |

### Analysis of the Hypotheses

Eye-movement data was used to assess differences in attentional engagement and change of focus between participants with and without chronic pain (H1). To conduct this assessment, we examined differences in *fixation duration*, *saccade frequency*, and *visit duration* between participants with and without chronic pain for the Questions and Answer AOIs on each of the three visual stimulus (*pain interference*, *physical function*, and *pain intensity*).

From the self-reported ratings, we used only the one that captured pain intensity. This is because we needed participants’ ratings for this measure to investigate a possible association between their objective eye movement data and their subjective pain intensity scores (H2). To examine this possibility, we conducted a backward regression utilizing the following equation for each stimulus:

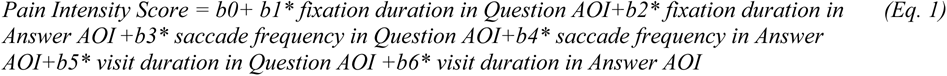

## Result

In this section, we report the results of our hypotheses. We start by reporting the results of hypothesis 1 and then discuss the results of hypothesis 2 for each of the three visual stimuli used in our study, pain interference, physical function, and pain intensity.

### Hypothesis 1

#### Pain Interference Stimulus

The comparison of fixation duration, saccade frequency, and visit duration in the Answer and Question AOI between the two groups (chronic pain, pain free) for the pain interference stimulus is shown in Figure 3. Participants in the chronic-pain group exhibited significantly (p=0.00) shorter fixation duration (52%) in the Question AOI than participants in the pain-free group (61%); significantly (p=0.04) fewer saccadic eye movements (60%) in the Question AOI than participants in the pain-free group (68%); and significantly (p=0.02) shorter visit duration (49%) in the Question AOI than the pain-free group (58%).

**Figure 3.**
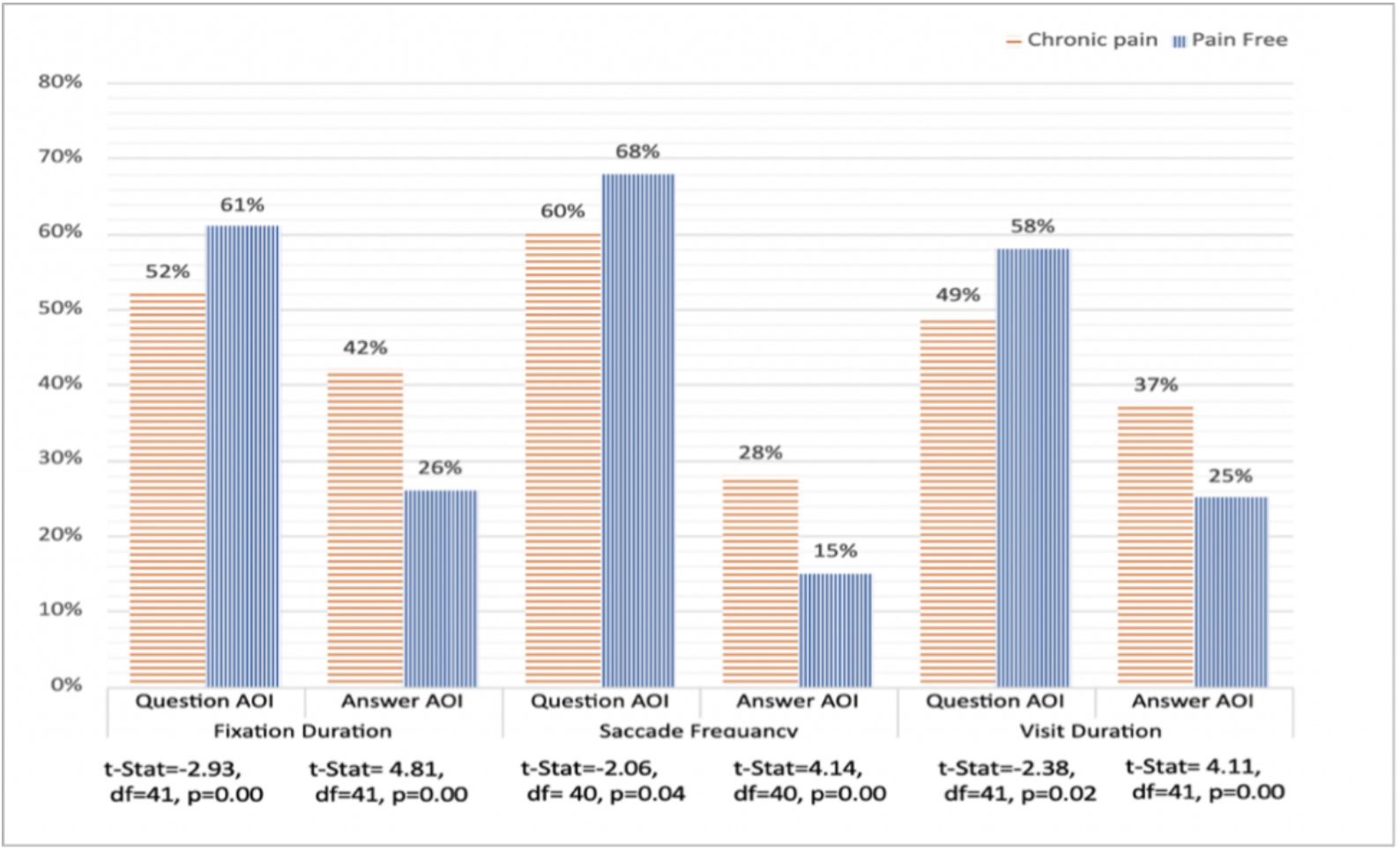
t-test results for H1 for the pain interference stimulus

The viewing behavior observed in Question AOI was reversed in Answer AOI. People in the chronic-pain group had significantly (p=0.00) longer fixation duration (42%) in the Answer AOI than participants in the pain-free group (26%); significantly (p=0.00) more saccades (28%) in the Answer AOI than the pain-free group (15%); and significantly longer visit duration (37%) in the Answer AOI than the pain-free group (25%).

In summary, the above results showing significant differences in engagement (fixation and visit duration) and change in focus (saccades) between the two groups support our first hypothesis for the pain interference stimulus.

#### Physical Function Stimulus

Similar to viewing trends that we detected in the pain interference stimulus; we observed reverse patterns of behavior between the two groups in Question and Answer AOIs of the physical function stimulus. While compared to the pain-free group, people in the chronic pain group showed less intense engagement and change of focus in the Question AOI, they exhibited more intense viewing behavior than the pain-free group in the Answer AOI. Our results showed that the chronic-pain group had significantly (p=0.03) shorter fixation duration (43%) in the Question AOI than the pain-free group (48%); had significantly (p=0.02) fewer saccadic eye movements (33%) than the pain-free group (53%) in the Question AOI; and had significantly (p=0.04) shorter visit duration in the Question AOI (41%) than pain-free group (46%) (Figure 4).

**Figure 4.**
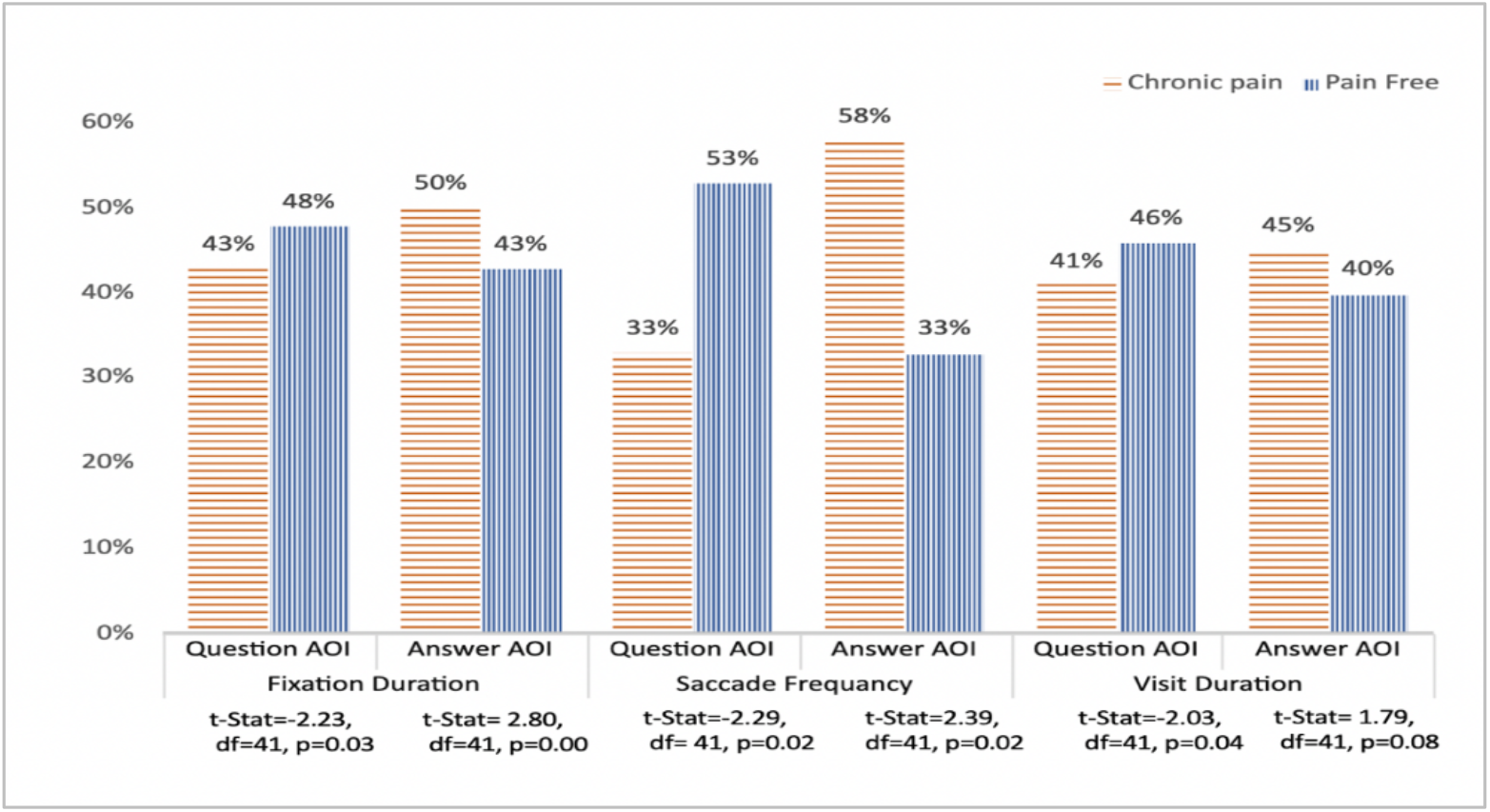
t-test results for H1 for the physical function stimulus

Fixation duration in the Answer AOI was significantly (p=0.00) longer for the chronic pain group (50% vs 43%). Also, the chronic pain group had significantly (p=0.02) more saccadic eye movement in the Answer AOI (58% vs 33%). The same pattern of behavior also was observed for visit duration in the Answer AOI. The chronic pain group had longer visit duration (45%) than the pain-free group (40%); however, the result was not significant at p=0.05 level (0.08).

Our analysis shows significant differences in engagement and change of focus between people with and without chronic pain. Hence, our analysis supports H1 for the physical function stimulus.

#### Pain Intensity Stimulus

Figure 5 shows the differences in fixation duration, number of saccades, and visit duration in each AOI between the two groups. Unlike the patterns observed in previous stimuli (pain interference and physical function), participants in the chronic pain group demonstrated longer fixation durations, more frequent saccadic eye movements, and longer visit durations in both Question- and-Answer AOIs than participants in the pain-free group. The differences between the two groups, however, were not significant. Hence our first hypothesis was not supported for the pain intensity visual stimulus.

**Figure 5.**
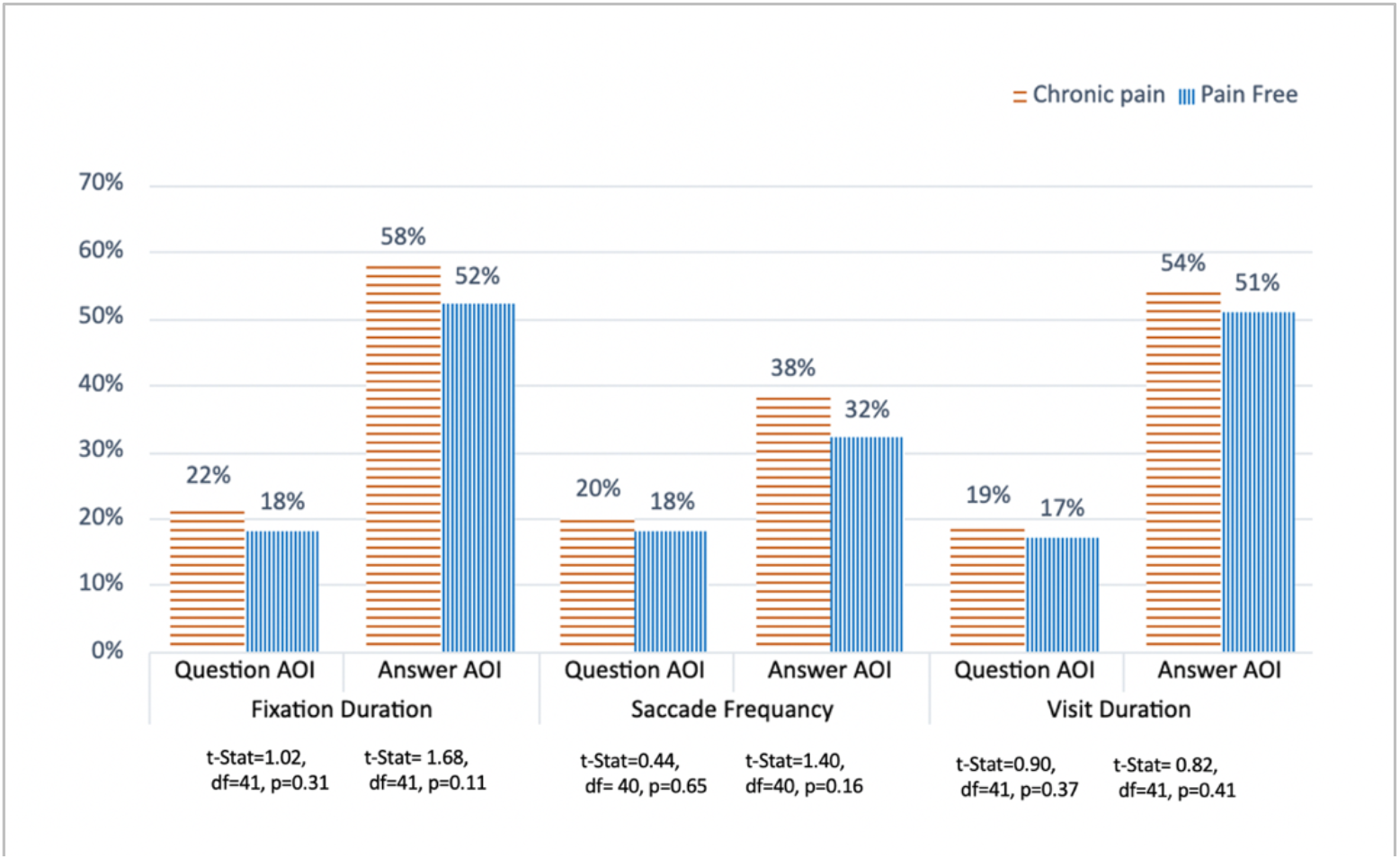
t-test results for H1 for the pain intensity stimulus

### Hypothesis 2

#### Pain Interference Stimulus

To assess the potential for fixation duration, frequency of saccades, and visit duration in the Question and Answer AOIs in the pain interference stimulus to predict the self-reported pain intensity scores, we conducted a backward regression analysis. Table 3 displays the results of the last step of the backward regression analysis, where the independent variable with the largest p-value was removed in each iterative step. The findings in Table 3 reveal a robust positive association between objective eye movements and subjective pain-intensity scores. Specifically, the results show that participants’ fixation duration in both AOIs and number of saccadic eye movements in the Answer AOI explained 51% of the variance in self-reported pain intensity ratings. This positive association between the dependent and independent variables indicates the longer fixation duration (engagement) in both AOIs and the more saccadic eye movements (change in focus) in the Answer AOI the higher the pain intensity score (p *<0.001*). These results show that participants’ eye movements indicating engagement and change of focus predicted their subjective pain experience. Hence, these results support H2 for the pain interference stimulus.

**Table 3:**
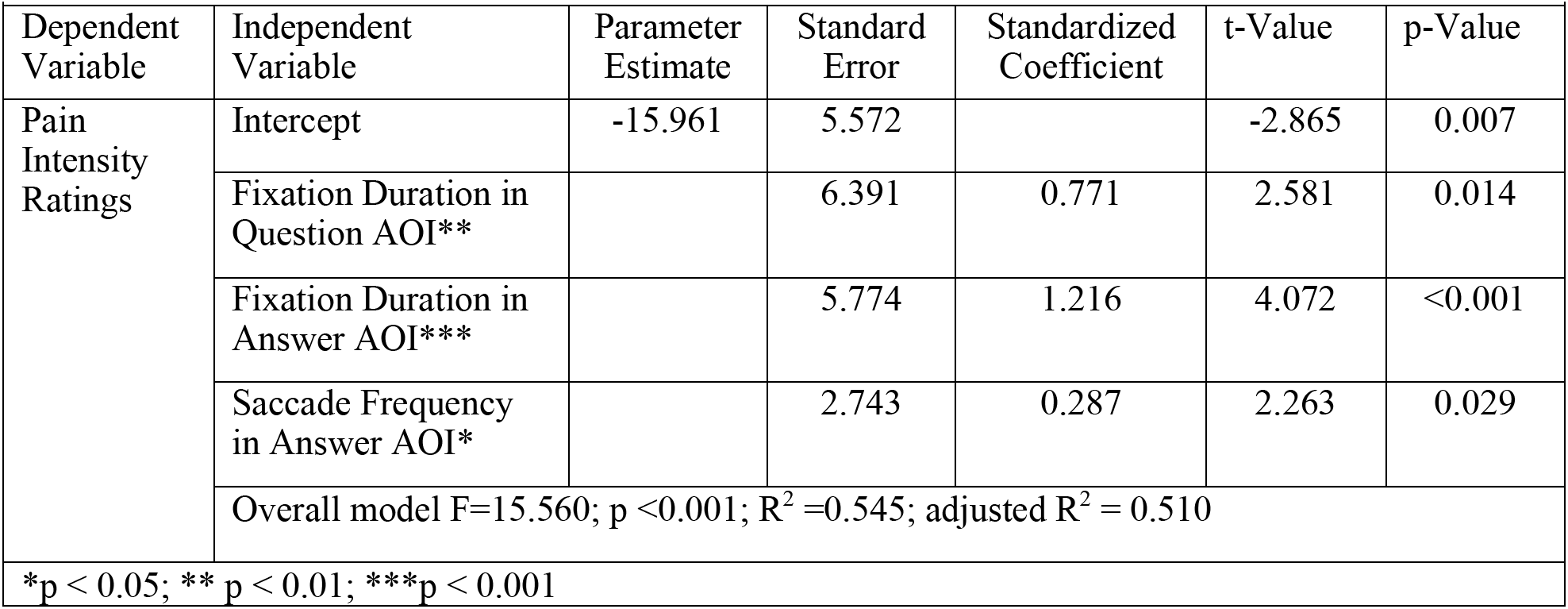
Regression result for H2 for the pain interference stimulus.

| Table 3: Regression result for H2 for the pain interference stimulus |  |  |  |  |  |  |
| --- | --- | --- | --- | --- | --- | --- |
| Dependent Variable | Independent Variable | Parameter Estimate | Standard Error | Standardized Coefficient | t-Value | p-Value |
| Pain Intensity Ratings | Intercept | -15.961 | 5.572 |  | -2.865 | 0.007 |
|  | Fixation Duration in Question AOI** |  | 6.391 | 0.771 | 2.581 | 0.014 |
|  | Fixation Duration in Answer AOI*** |  | 5.774 | 1.216 | 4.072 | <0.001 |
|  | Saccade Frequency in Answer AOI* |  | 2.743 | 0.287 | 2.263 | 0.029 |
|  | Overall model F=15.560; p <0.001; R<sup>2</sup>=0.545; adjusted R<sup>2</sup> = 0.510 |  |  |  |  |  |
| *p < 0.05; ** p < 0.01; ***p < 0.001 |  |  |  |  |  |  |

#### Physical Function Stimulus

The results of the backward regression for the physical function stimulus showed a strong inverse relationship between fixation duration in the Question AOI and the subjective pain intensity scores. That is, the shorter the fixation duration when reading the questions, the larger the value of pain intensity (Table 4). The results showed that 12% of the variance in subjective pain intensity scores was explained by fixation duration in the Question AOI (p=0.013). These results showed that a participant’s engagement (fixation duration) with the content of the Question AOI predicted the participant’s subjective pain intensity rating. While these results support H2, the R^2^ for the physical function stimuli (Table 4) is lower than the R^2^ for the pain interference stimuli (Table 3) reported in the previous section.

**Table 4:** Regression result for H2 for the physical function stimulus.

| Table 4: Regression result for H2 for the physical function stimulus |  |  |  |  |  |  |
| --- | --- | --- | --- | --- | --- | --- |
| Dependent Variable | Independent Variable | Parameter Estimate | Standard Error | Standardized coefficient | t-Value | p-Value |
| Pain Intensity Ratings | Intercept | 7.764 | 1.933 |  | 4.017 | <0.001 |
|  | Fixation Duration in Question AOI** | -10.638 | 4.119 | -0.0374 | -2.583 | 0.013 |
|  | Overall model F= 6.670; p <0.010; R<sup>2</sup> =0.140; adjusted R<sup>2</sup> = 0.119 |  |  |  |  |  |
| *p < 0.05; ** p < 0.01; ***p < 0.001 |  |  |  |  |  |  |

#### Pain Intensity Stimulus

Table 5 displays the last step of the backward regression analysis that was conducted to examine the association between objective eye-movement behaviors and subjective pain intensity ratings for the pain intensity stimulus. As shown in this table, the p-value for the independent variable, while close to the accepted threshold, does not reach the required 0.05 value. Hence, this analysis shows that eye movements representing engagement (fixation and visit duration) and change of focus (saccade frequency) on this visual stimulus did not predict participants’ subjective pain intensity ratings. In other words, H2 was not supported for this visual stimulus.

**Table 5:** Regression result for H2 for the pain intensity stimuli.

| Table 5: Regression result for H2 for the pain intensity stimuli |  |  |  |  |  |  |
| --- | --- | --- | --- | --- | --- | --- |
| Dependent Variable | Independent Variable | Parameter Estimate | Standard Error | Standardized coefficient | t-Value | p-Value |
| Pain Intensity Ratings | Intercept | -0.232 | 1.627 |  | -0.143 | .887 |
|  | Fixation Duration in Answer AOI | 5.580 | 2.858 | 0.292 | 1.950 | 0.058 |
|  | Overall model F= 3.81; p = 0.058; R<sup>2</sup>=0.085; adjusted R<sup>2</sup> = 0.063 |  |  |  |  |  |
| *p < 0.05; ** p < 0.01; ***p < 0.001 |  |  |  |  |  |  |

### Hypothesis Testing Summary

Table 6 summarizes the results of the study. As shown in Table 6 hypotheses 1 and 2 are supported for two of the three visual stimuli used in our study, namely pain interference and physical function.

**Table 6:** Summary of results.

| Table 6: Summary of results |  |  |  |  |  |  |  |
| --- | --- | --- | --- | --- | --- | --- | --- |
| *H1. Completing pain-related surveys provides a sensitive stimuli-task paradigm for capturing differences in attentional bias between people with and without chronic pain.* |  |  |  |  |  |  |  |
| Stimulus | Q_AOI |  |  | A_AOI |  |  |  |
|  | FD | SF | VD | FD | SF | VD |  |
| Pain interference | †CP<PF | †CP<PF | †CP<PF | †CP>PF | †CP>PF | †CP>PF | Supported |
| Physical function | †CP<PF | †CP<PF | †CP<PF | †CP>PF | †CP>PF | CP>PF | Supported |
| Pain intensity | CP>PF | CP>PF | CP>PF | CP>PF | CP>PF | CP>PF | Not Supported |
| *H2. Completing pain-related surveys provides a sensitive stimuli-task paradigm for predicting subjective pain intensity scores from objective eye movements:* |  |  |  |  |  |  |  |
| $PIS = b_0 + b_1 * FD_{Q\_AOI} + b_2 * FD_{A\_AOI} + b_3 * SF_{Q\_AOI} + b_4 * SF_{A\_AOI} + b_5 * VD_{Q\_AOI} + b_6 * VD_{A\_AOI}$ | | | | | | | |
| Pain interference | $PIS = b_0 + b_1 * FD_{Q\_AOI} + b_2 * FD_{A\_AOI} + b_3 * SF_{A\_AOI}$ $^{\dagger}F = 15.560; p < 0.001; R^2 = 0.545; \text{adjusted } R^2 = 0.510$ | | | | | | Supported |
| Physical function | $PIS = b_0 + b_1 * FD_{Q\_AOI}$ $^{\dagger}F = 6.670; p < 0.010; R^2 = 0.140; \text{adjusted } R^2 = 0.119$ | | | | | | Supported |
| Pain intensity | $PIS = b_0 + b_1 * FD_{A\_AOI}$ $F = 3.810; p = 0.058; R^2 = 0.085; \text{adjusted } R^2 = 0.063$ | | | | | | Not Supported |
| Q_AOI= Question AOI; A_AOI= Answer AOI; FD=fixation duration; SF=saccade frequency; VD= visit duration; CP= chronic pain; PF=pain free; PIS=pain intensity score; †=significant results |  |  |  |  |  |  |  |

## Discussion

While the literature provides ample evidence that pain impacts cognition, it suggests that the ability to capture attentional biases that are caused by the interruptive function of pain are largely dependent on stimuli-task paradigms (Chan et al., 2020). Grounded in the chronic-pain literature, we argued that stimuli-task paradigms appropriate for the NeuroIS research that focuses on designing smart clinician support systems for chronic pain are (1) context-rich and pain-related, (2) engage people directly in appraising their pain experience, and (3) do not impose short exposure time limits. Stimuli-task paradigms that meet these criteria are likely to provide more opportunities for capturing the impact of chronic pain on visual attention. Hence, they are likely to be effective in detecting differences in information-processing behavior between people with and without chronic pain.

To test this argument, we used a task that engaged our study participants directly in appraising their pain experience by asking them to complete three pain-related measures in the PROMIS 29+ profile (i.e., pain interference, physical function, and pain intensity) that required them to summarize their pain-related health symptoms in the past seven days. These three pain-related surveys served as the visual stimuli for the task. We collected participants’ eye movements as they completed these surveys.

To prepare the collected data for hypothesis testing, we classified the eye-tracking datasets into two distinct groups based on participants’ self-identification as belonging to either the chronic-pain or pain-free groups, based on whether they experienced persistent pain for at least three months. We hypothesized that using our proposed stimuli-task paradigm we would be able to detect significant disparities in visual behavior between the chronic-pain and pain-free groups when they completed the experimental task of answering the survey questions.

To investigate the differences in visual behavior, we compared ***engagement with the stimuli*** and the ***frequency of changes in focus (attentional shifts)*** between the two groups, with engagement and change of focus being two commonly used visual behaviors in the literature for assessing the impact of pain on visual attention (Fashler and Katz, 2014, 2016; Skaramagkas et al., 2023; Yang et al., 2012). The differences in information processing behavior between the two groups were examined for two relevant AOIs on each stimulus: Question AOIs that covered the question area and Answer AOIs that covered responses to questions on visual stimuli used in our study (Figure 1). Consistent with previous pain research, ***differences in engagement*** between the two groups were determined by examining ***fixation*** and ***visit duration*** (Alrefaei et al., 2022; Fashler and Katz 2014, 2016; Vervoort et al., 2013), while ***differences in change of focus*** were determined by analyzing ***saccade frequency*** (Skaramagkas et al., 2023).

Hypothesis one focused on examining whether our proposed stimuli-task paradigm was sensitive in detecting differences in viewing behavior between people with and without chronic pain. Our analysis confirmed this hypothesis for two of the three visual stimuli used in our study. Specifically, the results revealed significant differences in how the two groups processed information on the ***pain interference*** and ***physical function*** stimuli but not on the ***pain intensity*** stimulus. One potential explanation for this discrepancy is that the pain intensity stimulus was less contextually rich compared to the other two stimuli, as it featured only one item (as compared to the other two stimuli that had 4 items each). This interpretation aligns with the notion that the more contextually elaborate the stimuli, the greater the prospects for capturing nuances in visual attention.

The detected differences in information processing behavior for both AOIs of the pain interference and physical function stimuli revealed an interesting trend. People in the chronic-pain group exhibited significantly less cognitive effort (e.g., less intense engagement and fewer changes in focus) than people in the pain-free group when reading the questions. When it came to responding to questions, however, they expended significantly more cognitive effort than people in the pain-free group. The fact that people in chronic pain expended more cognitive effort when responding to questions is not surprising. Naturally, people with chronic pain, compared to those who are pain free, have more pain-related incidences to recall and evaluate (Alrefaei et al., 2022). Paying less attention to questions (compared to those who were pain free) may be due to avoidance behavior. According to pain literature, vigilance/avoidance behavior toward pain-related stimuli is determined by level of pain intensity. When pain is severe people exhibit vigilance and when pain is not severe, they exhibit avoidance behavior (Eccleston and Crombez, 1999). To provide insight into pain intensity levels of participants in the chronic pain group, we examined the participants’ ratings for the PROMIS 29+ v2 pain intensity measure. The measurement of subjective pain intensity is conducted through a numeric rating scale consisting of 11 points, ranging from 0 (no pain) to 10 (worst pain imaginable). Ratings in 7 to 10 range indicate severe pain (Karcioglu et al., 2018). The pain intensity scores in our study show that our participants in the chronic-pain group did not suffer from severe pain (M = 5.095, SD= 1.410). Hence, our results support the literature that suggests when pain intensity is not severe, people may engage in avoidance behavior. That is, people in the chronic-pain group may have focused on completing the task (e.g., accurately summarizing pain experience in a single score) to escape from negative thoughts and feelings that were invoked by reading pain-related questions (Eccleston and Crombez, 1999).

Hypothesis two tested the sensitivity of our stimuli-task paradigm in detecting a possible relationship between participants’ subjective pain intensity scores and their objective visual behavior. Specifically, we sought to determine whether eye movements that reflect engagement and change in focus could predict individuals’ self-reported pain intensity scores. Our findings revealed significant associations between participants’ objective attentional eye-movement behavior, specifically engagement and change in focus, and their subjective pain-intensity score for the pain interference and physical function stimuli. In contrast, no significant associations were observed for the pain intensity visual stimuli (the third stimulus). These outcomes are in line with our hypothesis 1 testing results, which underscores the notion that stimuli with more contextual information, such as pain interference and physical function, have greater potential to reveal nuances of attention. Similarly, the obtained results for H2 in our study indicate that context-rich stimuli are likely to be more effective in predicting subjective pain intensity scores compared to those with fewer contextual details.

Our results show that the first stimulus, ***pain interference***, was the most sensitive visual stimuli in our study for detecting differences in engagement and change in focus between participants with and without chronic pain. The differences between the two groups were significant for both engagement and change in focus in both AOIs for this stimulus. Similarly, 51% of the variance in subjective pain intensity scores was explained by visual behavior on pain interference stimulus. For the second stimulus, ***physical function***, all but one t-test showed significant differences between the two groups; visual engagement (measured as visit duration) between the two groups in the Answer AOI did not reach significance at the 0.05 level. While the association between objective eye movements and subjective pain intensity scores were significant, this relationship was weaker for the physical function stimulus compared to the one observed for the pain interference stimulus (12% vs. 51%).

One reason could be that participants in our study found the items in the pain interference stimuli more relevant than those in the physical function stimuli (Chan et al. 2020). The former focused more broadly on assessing pain interference with daily activities (i.e., “How much did pain interfere with your day to day activities?”, “How much did pain interfere with work around the home?”, “How much did pain interfere with your ability to participate in social activities?”, and “How much did pain interfere with your household chores?”). The latter focused on the impact of pain on relatively more specific physical daily activities (i.e., “Are you able to do chores such as vacuuming or yard work?”, “Are you able to go up and down stairs at a normal pace?”, “Are you able to go for a walk of at least 15 minutes?”, and “Are you able to run errands and shop?”).

Our participants in the chronic-pain group suffered from a diverse range of pain experiences. As a result, the pain questions may not have been relevant to everyone in pain. Questions in the ***physical function stimuli*** such as “Are you able to go up and down stairs at a normal pace”, or “Are you able to go for a walk of at least 15 minutes?” maybe more relevant to those who suffer from back, leg, muscle, and knee pain compared to those who suffer from headache, wrist, shoulder, and eye pain (Chan et al., 2020). Questions in the ***pain interference stimuli***, however, seemed to be relevant to all the reported chronic conditions in our study. Future research is needed to examine this possibility.

## Contributions and Implications

Our study makes important contributions to NeuroIS literature by proposing and testing a new stimuli-task paradigm that provides more opportunities for capturing differences in visual behavior between those who suffer from chronic pain and those who are pain free. In particular, our investigation showed that two of the three visual stimuli used in our study were not only sensitive in revealing differences in eye movements that represent engagement and change in focus between people with and without chronic pain, but also sensitive in facilitating the predictive capacity of these two eye movement behaviors to forecast a participant’s pain intensity score regardless of whether the participant self-identified as someone who suffers from chronic pain or someone who is pain free.

We calculated three eye-tracking metrics (fixation duration, saccade frequency, and visit duration) to measure information processing behavior in our study. We normalized these eye-tracking metrics by calculating the ratios of their total values (Shojaeizadeh et al., 2019). Such normalization is often important for stimuli-task paradigms that are not completed in fixed time intervals (e.g., dot-probe tasks). Our results show that the calculated eye-tracking metrics used in our study were effective in capturing differences in information-processing behavior between people with and without chronic pain. Our results also showed a strong association between participants’ objective visual attention (captured by the eye-tracking metrics used in our study) and their subjective pain scores. By doing so, our study provides support and rationale for NeuroIS research that focuses on developing smart information systems for detecting chronic pain experience from eye movement data.

Our study also contributes to pain literature. Our results suggest that eye movements have the potential to serve as reliable biomarkers of chronic pain. Our study extends the existing stimuli-task paradigms for investigating the interruptive function of pain on attention to include pain-related surveys as visual stimuli, and survey completion as the experimental task. Our results also show that the eye-tracking variables that we developed for our proposed paradigm (i.e., FD, SF, VD in Question and Answer AOIs as ratios of their total values) were effective in detecting the impact of chronic pain on visual attention. By offering more opportunities for capturing nuances in information processing and decision behavior, and eye-tracking variables that can effectively capture such nuances, our proposed stimuli-task paradigm is likely to assist pain researchers in exploring and explaining how pain disrupts attention.

The practical implications of our study hold significant potential for enhancing the assessment of chronic pain in clinical settings. Pain-related measures in the PROMIS profile are commonly utilized in clinical settings to assess pain. By using response to these measures as a stimuli-task paradigm, our results demonstrate that both objective eye movements and subjective ratings can be collected via smart information systems, thus enabling the collection of data in advance of routine office visits in hospital and office settings. Modern eye-tracking devices can be readily integrated with information systems administering pain surveys in clinical settings, thereby facilitating the acquisition of high-quality eye movement data. The increasing integration of high-quality eye-tracking devices in consumer-grade computing products represents a promising trend, offering the potential for remote collection of eye movement data during virtual office visits. These developments hold promise for the development of smart clinician support systems that can support the advancement of chronic pain management, hence, further underscore the importance of continued research in this domain.

## Limitations and Future Research

As with any experiment, our study has strengths and limitations. One important strength in our study is that we used a relatively more diverse population than previous eye-tracking studies by recruiting participants from within and outside the university campus (e.g., Alrefaei et al., 2022). Having a diverse set of chronic pain types present in our study could be considered as both strength and limitation. On one hand, our results showing that our proposed stimuli-task paradigm and eye-tracking variables are effective in detecting the differences in information processing behavior between people with and without chronic pain regardless of their pain types, attests to the robustness of our proposed stimuli-task paradigm and eye-tracking variables. On the other hand, focusing on visual behavior of a population that suffers from a specific chronic pain may enable us detect nuances in information processing behavior that may be unique to that particular chronic pain condition. Future research is needed to provide insight into the above-stated interpretations.

Using only three visual stimuli in our study could be viewed as a limitation. The significant results in our study, however, with only three stimuli show that we may be able to detect differences in visual behavior between people with and without chronic pain by using even fewer stimuli, e.g., only one 4-item subjective measure that requires people to appraise their pain symptoms on a 5-point scale (e.g., pain interference or physical function).

Similarly, using only three eye-tracking variables (fixation duration, saccade frequency, and visit duration) to investigate attention and change in focus could be considered as a limitation. Our results, however, conclusively demonstrate the capacity of these three variables to successfully detect changes in the visual behavior between those who suffer from chronic pain and those who are pain free. Nevertheless, future studies with different pain-related surveys and eye-tracking variables are needed to verify and extend our results.

One of the key aims of our study was to demonstrate that chronic pain status and the intensity of pain can be accurately detected solely through the analysis of eye movements. Recent studies show that other objective measures such as mouse movements can reveal users experience of cognitive load and cognitive deliberation when they complete online forms (Weinmann et al. 2021; Kim et al. 2022). Future studies can extend our work by combining eye movements with mouse movements to study how chronic pain impacts information-processing behavior.

## Conclusion

The pressing need for objective measures in the evaluation of chronic pain both in research and practical contexts (Borsook et al., 2011) underscores the significance and value of NeuorIS research in this pivotal domain of human health. Our study shows that subjective pain-related measures that are now commonly used in clinical settings can serve as effective visual stimuli for detecting marked differences in ocular behavior of individuals with and without chronic pain and for predicting how an individual would rate his/her pain intensity.

Our study also shows that our eye-tracking variables were effective in measuring the ocular behavior for the proposed stimuli-task paradigm. These findings underscore the potential value of NeuroIS research in identifying practical stimuli-task paradigms that can be implemented in clinical settings. They also highlight the value of NeuorIS research in determining eye movement metrics that can serve as reliable predictors (i.e., biomarkers) of chronic pain. Such research endeavors have the potential to foster the development of cutting-edge machine learning algorithms and intelligent clinician support systems that can detect chronic pain experience automatically, and thereby assist clinicians in enhancing the quality of patient care.

## Data Availability

All data produced in the present work not available to share.

## Appendix A

The results of a recent SLR paper (Chan et al. 2020) highlight the paucity of eye-tracking studies that investigate the impact of chronic pain on attentional processes. The review article found only 24 studies that used eye tracking to examine how pain-related stimuli impact attentional processes. Only 11 papers among the 24 reviewed articles investigated the interruptive function of chronic pain on attention. Among the 11 papers that investigated chronic pain, only 9 included both chronic pain and healthy participants. Because our study examines the differences in ocular behavior of people with and without chronic pain, we summarized these 9 reviewed articles in Table 7. The complete details of all 24 reviewed articles are provided in the SLR paper (Chan et al. 2020, Table A1, pp. 5-10).

**Table 7:**
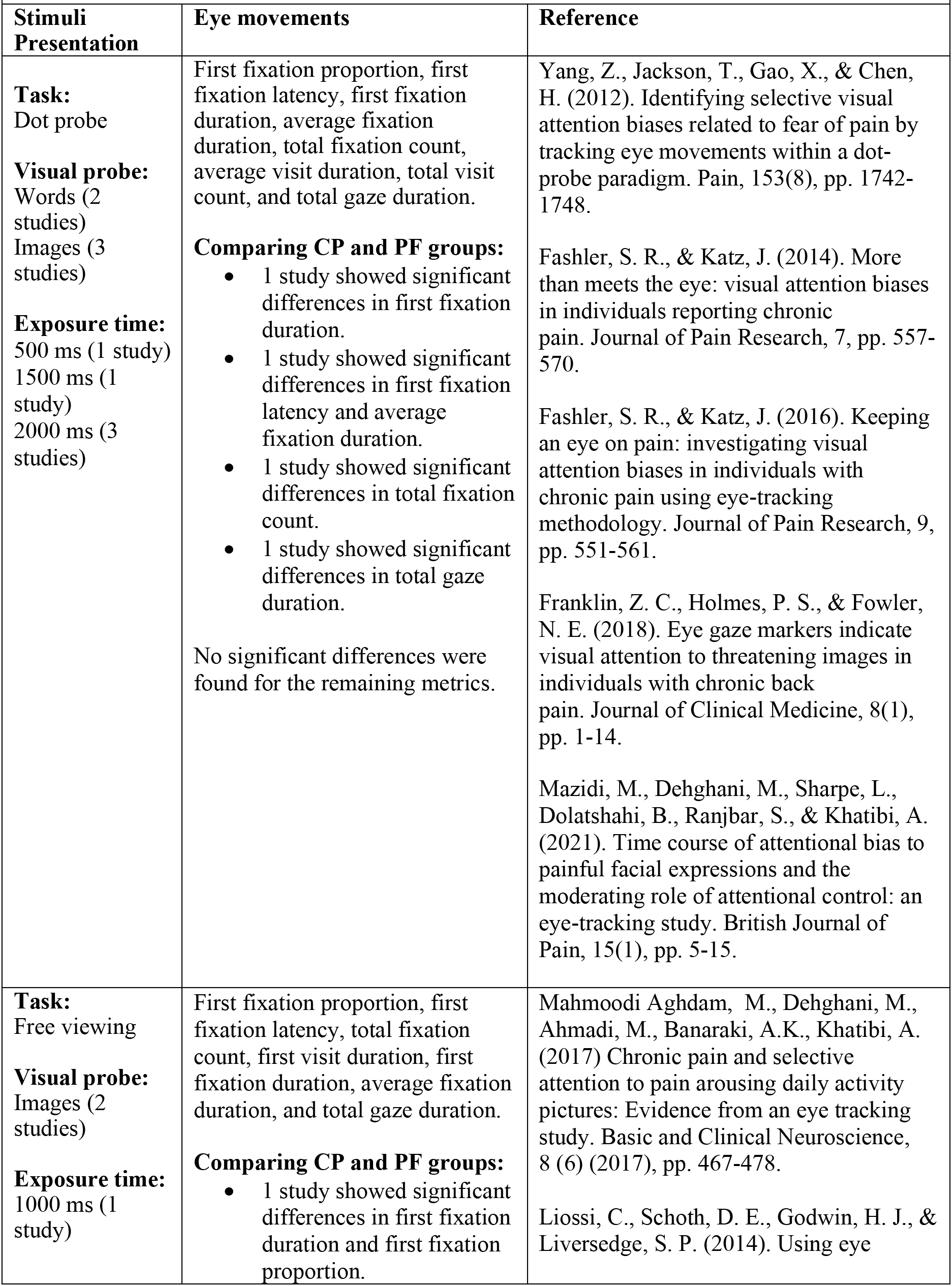
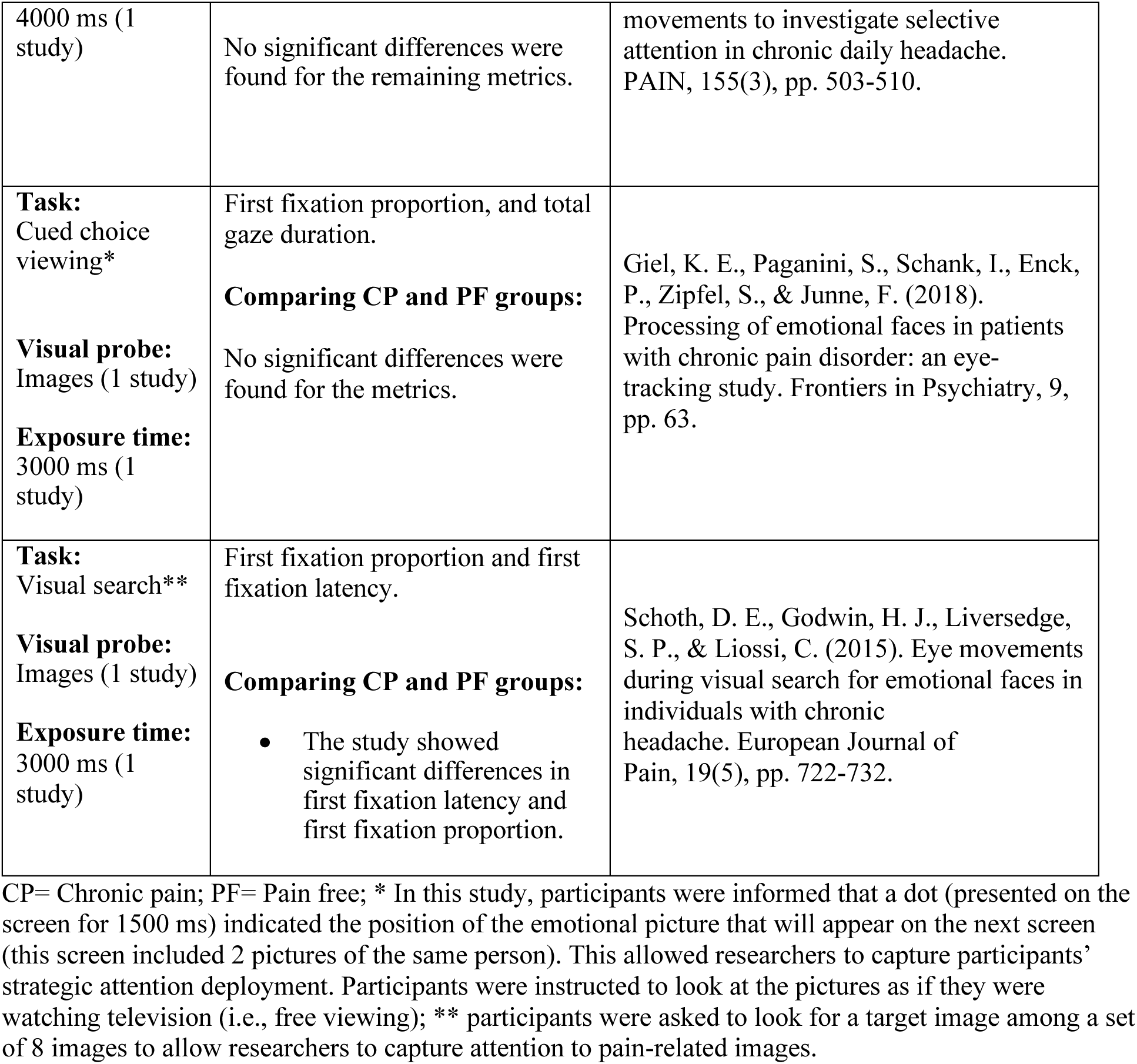
Major characteristics of the stimulus presentation paradigm and their related eye-movement measures that were used in eye-tracking chronic pain studies that compared the ocular behavior of people with and without chronic pain.

The 9 studies that are summarized in Table 7, all use short, fixed exposure times (500-4000 ms) to capture reaction to pain-related words or images. Dot-probe is the most frequently used task in these 9 studies.

The findings reported in Table 7 show mixed results. For example, *first fixation duration* was significantly different between people with and without chronic pain in a study that used the dot-probe task and a study that used the free viewing task. The same eye-movement metrics, however, did not show significant differences between chronic pain and healthy individuals in five other studies that used either the dot-probe task or the free viewing task. Similarly, *first fixation latency* was significantly different between people with and without chronic pain in two studies (one used the dot-probe task and the other used the visual search task) but not in other studies that used similar tasks.

## Notes

### Competing Interest Statement

The authors have declared no competing interest.

### Funding Statement

This study did not receive any funding

### Author Declarations

Ethics committee/IRB of Worcester Polytechnic Institute gave ethical approval for this work

